# Interspecies normalization of dose response relationship for adeno-associated virus-mediated hemophilia gene therapy – application to first-in-human dose prediction

**DOI:** 10.1101/2022.07.09.22277452

**Authors:** Peng Zou

## Abstract

Establishing dose response relationships in targeted patients is foundational in the development of therapeutic drugs including gene therapy products. Enlightened by interspecies normalization of plasma drug concentration-time curves using Dedrick plot, the author of this manuscript first demonstrated the feasibility of normalizing dose-response relationship of adeno-associated virus (AAV)-mediated hemophilia gene therapy products in multiple species to a species-invariant scale. Preclinical dose-response relationships of eight AAV vectors were normalized using an exponent of 0.25 and applied to first-in-human (FIH) dose prediction. The performance of this dose-response normalization approach for FIH dose prediction was compared to that of direct body weight-based dose conversion and allometric scaling approaches. The study results suggested that in addition to hemophilia dogs and non-human primates, inclusion of larger animal models (e.g., swine and cattle) in preclinical dose-finding studies of AAV vectors might improve the performance of interspecies dose-response normalization approach. Furthermore, it was found that AAV capsid-specific T cell responses in hemophilia patients might cause underprediction of FIH dose while novel bioengineered capsids with a high transduction efficiency specifically in human hepatocytes might cause overprediction of FIH dose. These factors should be considered when dose-response is extrapolated from preclinical species to patients.

## Introduction

With the approvals of Luxturna® and Zolgensma® in Europe and the United States, recombinant adeno-associated virus (rAAV)-based gene therapies (GT) have shown promise for the treatment for diseases with genetic disorders. Establishing dose response relationships in targeted patients is foundational in the development of therapeutic drugs including GT. The efficacy and safety of adeno-associated virus (AAV)-based GT are largely dose-dependent in both preclinical species and humans.^1^ However, it is still challenging to extrapolate dose response of GT from animals to humans. For example, dose response curves in preclinical species and humans are usually nonlinear and does not follow a consistent pattern among species.^2^ Traditional quantitative methods such as PK-PD modeling used to predict human dose-response relationships of small molecules and therapeutic proteins are difficult to apply to GT, and model structures and parameter paradigms may not be directly applicable to these complex GT products.^3^ A mechanistic modeling approach incorporated with disposition of vector in the body, transduction efficiency in target tissues, expression strength in transduced cells, and duration of expression has been proposed to establish human dose response relationships for GT.^2^ However, our current understanding of GT pharmacology is still limited, and experimental data are usually inadequate to validate a complex mathematical model for GT products especially at early development stage.

It was reported that the metabolic rate of cells (B_c_) in vivo in mammalian species decreased with increasing body weight (B_c_ = B_0_ / W^0.25^, where B_0_ is the cellular metabolic rate expressed as 3 × 10^−11^ Watts per cell for organism with mass of 1.0 gram, and W is the mass of a mammalian species).^4^ Since intracellular synthesis of transgene protein following GT is dependent on metabolic rate of cells, Tang et. al. proposed a concept of gene efficiency factor (GEF) to describe the efficiency of gene transfer system and demonstrated a linear relationship between logGEF and logW using preclinical and clinical data of three AAV vectors for hemophilia B therapy, where W was the body weight of mammalian species.^5^ Tang et. al. concluded that body weight-based cross-species allometric scaling of GEF could be used to predict first-in-human (FIH) dose of AAV-mediated hemophilia B GT. However, Aksenov et. al. pointed out the limitation of Tang’s allometric scaling approach that Tang et. al. assumed a linear dose response relationship for serum factor IX (FIX) while the dose-response curves observed in most preclinical species and hemophilia B patients were nonlinear.^2^ Then, Aksenov et. al. used a powder regression model (FIX concentration = a × Dose^b^) to describe dose-FIX concentration relationships for AAV-FIX vectors, where a was proportionality coefficient and b was the exponent for dose. Aksenov et. al.’s analysis showed that the dose response curves of AAV vectors did not follow a consistent pattern across species and no obvious relationship for proportionality coefficient or exponent vs. body weight was observed. Therefore, Aksenov et. al. concluded that Tang’s allometric scaling was unable to accurately predict human dose for AAV GT.

Enlightened by interspecies normalization of plasma drug concentration-time curves using Dedrick plot,^6^ this author hypothesized that interspecies normalization approach might be applied to dose-response extrapolation from animals to humans. The Detrick plot approach, also called species-invariant time method, assumes that by normalizing the concentrations with body weight and transforming chronological time to physiological time, the plasma drug concentration–time curves should be superimposable in all species.^7^ The transformed concentration–time curves of various species are superimposed and then back-transformed to estimate human plasma concentration–time profile. Since the expression efficiency of a transgene product is reversely correlated with W^0.25^, for a transgene product that functions in systemic circulation, this author hypothesized that the total amount of transgene product in blood circulation across species could be normalized to a species-invariant scale using an exponent of 0.25. The normalized transgene product–dose curve can be back-transformed to predict human dose-response curve.

Among different types of AAV programs, hemophilia GT has more preclinical and clinical dose-response data available in literature. The most common types of hemophilia are hemophilia A and hemophilia B, caused by mutations in *F8* or *F9*, coding for factor VIII (FVIII) and factor IX (FIX) proteins, respectively.^8^ In this study, this author explored if the total serum amount of FVIII or FIX following AAV-mediated GT could be normalized across species and if the normalized dose response could be extrapolated to hemophilia patients. Furthermore, the normalized serum FVIII or FIX-dose curves were used to predict the FIH dose of AAV vectors. The predictive performance of this interspecies normalization approach was compared to that of two previously reported approaches, direct vg/kg conversion from preclinical doses and allometric scaling.

## Methods

### Data source

Preclinical and clinical transgene product expression data of vectors were collected from published literature. Only vectors with clinical data and preclinical data from ≥ 2 animal species available were collected. When the body weight of individual subject or animal was not available in literature, the body weights of mouse, cynomolgus macaque, rhesus macaque, and human were assumed as 0.02, 2.5, 8.0, and 70 kg, respectively.

### Targeted plasma transgene product levels

The FIH dose is anticipated to be effective in eliminating uncontrolled bleeds. Thus, 12% of normal plasma FVIII activity level and 5% of normal plasma FIX level were proposed as the targeted transgene protein levels for hemophilia A and B GT, respectively.^9^ The normal plasma level of FIX in healthy subjects is 5000 ng/mL ^10^ and thus a targeted FIX level of 250 ng/mL was used to project FIH doses of FIX vectors except for SPK-9001 vector encoding FIX-Pauda. FIX-Padua is a hyperfunctional variant of FIX and the residue 338 in human FIX is changed from arginine to alanine. FIX-Pauda encoded by SPK-9001 has an 8- to 12-fold increased specific activity (a mean difference of 9.1-fold in hemophilia B mice) compared to endogenous FIX.^11^ Therefore, the targeted plasma FIX-Pauda level was selected as 250 ng/mL ÷ 9.1 = 27.5 ng/mL (equivalent to 5% of normal FIX level).

### Interspecies normalization of dose-response and FIH dose prediction

Transgene product amount in the blood circulation of individual animal and human was normalized using Equation 1:

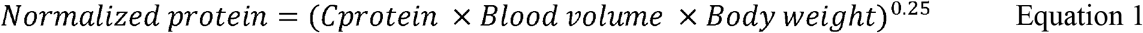

C_protein_ was the maximum plasma concentration of transgene product following GT. The blood volume of mouse, dog, cynomolgus macaque, rhesus macaque, and human was 79, 86, 65, 54, and 70 mL per kg body weight.^12^

A power regression model was used to correlate normalized transgene product amounts and the total vector doses (vg) received by different species. The regression was conducted using the data of three species (two animal species and human) and two animal species only. The targeted transgene product level (12 IU/dL for FVIII, 250 ng/mL for FIX, and 27.5 ng/mL for FIX-Pauda) was normalized using Equation 1. The normalized transgene product amount in patient’s blood circulation at the targeted level was then incorporated into the power equation derived from two preclinical species to calculate FIH dose.

### GEF calculation and allometric scaling

Individual animal and human GEF values were calculated using Equation 2 previously reported by Tang et. al..^5^

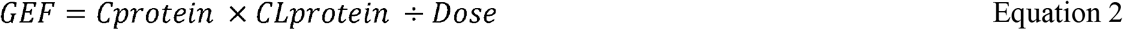

C_protein_ was the maximum plasma concentration (C_max_) of transgene product following GT. CL_protein_ was the total clearance of FIX or FVIII, which was the average clearance values from multiple animal/human pharmacokinetic studies (supplemental Table S1). Dose was the total vector genomes (vg) received by individual animal or patient. The mean GEF of each species was used for allometric scaling. A linear regression was conducted between logGEF and logW using the data of three species (two animal species and human) and two animal species only.^5^ The equation derived from two animal species was used to predict human GEF.

When the human GEF predicted by allometric scaling was available, Equation 3 was used to predict FIH dose.

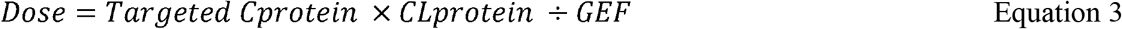

Where the targeted C_protein_ was 12 IU/dL for FVIII and 250 ng/mL for FIX (27.5 ng/mL for FIX-Pauda). GEF is the predicted human GEF derived from allometric scaling of two animal species.

### Direct vg/kg conversion

A linear regression was conducted between monkey or dog plasma FIX levels or FVIII activity levels at various doses and total vector doses (vg). Then, plasma FIX level of 250 ng/mL (27.5 ng/mL for FIX-Pauda) or plasma FVIII activity level of 12 IU/dL was incorporated into the regression equation to estimate monkey or dog total vector dose which was anticipated to produce transgene product in monkeys or dogs at targeted level. The total dose was divided by animal body weight and the weight normalized dose (vg/kg) was the FIH dose derived from the direct vg/kg conversion approach.

## Results

A total of five vectors for hemophilia A therapy and three vectors for hemophilia B therapy were included in this study. The rAAV2-CMV-FIX vector included in Tang et. al.’s report was excluded because plasma FIX levels were less than 1% of normal in hemophilia B patients receiving this vector across all dose levels and it was difficult to distinguish endogenous FIX and FIX produced by GT.^5^ The preclinical and clinical doses, plasma FVIII activity or FIX levels, normalized FIIIV or FIX in blood circulation, and calculated GEF for the 8 vectors were summarized in supplemental Tables S2 and S3. The data of individual animal or patient were listed in the tables if they were available in literature. Otherwise, only mean values were listed in the tables.

Normalized FIX or FVIII amounts in blood circulation of animals and individual patient were plotted against total vector doses (vg). A power regression model could describe the relationships between normalized transgene protein and dose across three species for the eight vectors (Figures 1 and 2). Four of the eight vectors showed three-species correlation R^2^ values > 0.8 and the other four vectors (scAAV2/8-LP1-hFIXco, GO-8, SB-525, and SPK-8011) showed three-species correlation R^2^ < 0.8. The low R^2^ values of the four vectors were mainly due to large inter-subject and inter-animal variability in transgene product levels and small sample size at each dose level. In most studies, AAV vectors were administered to less than six animals or patients at each dose level. When normalized mean FVIII or FIX amounts were used for regression, R^2^ values were substantially improved (supplemental Figures S1 and S2). Only scAAV2/8-LP1-hFIXco vector showed a three-species correlation R^2^ < 0.8 (R^2^ = 0.7682) (Figure S1C). The low R^2^ indicated a substantially lower gene expression activity of scAAV2/8-LP1-hFIXco in patients compared to animals, which was likely due to intense T cell responses to this vector in hemophilia B patients.^13, 14^ Overall, the moderate-to-strong correlation between normalized transgene product and vector dose (supplemental Figures S1 and S2) suggested that it was feasible to normalize dose response in multiple species to a species-invariant scale using an exponent of 0.25.

**Figure 1.**
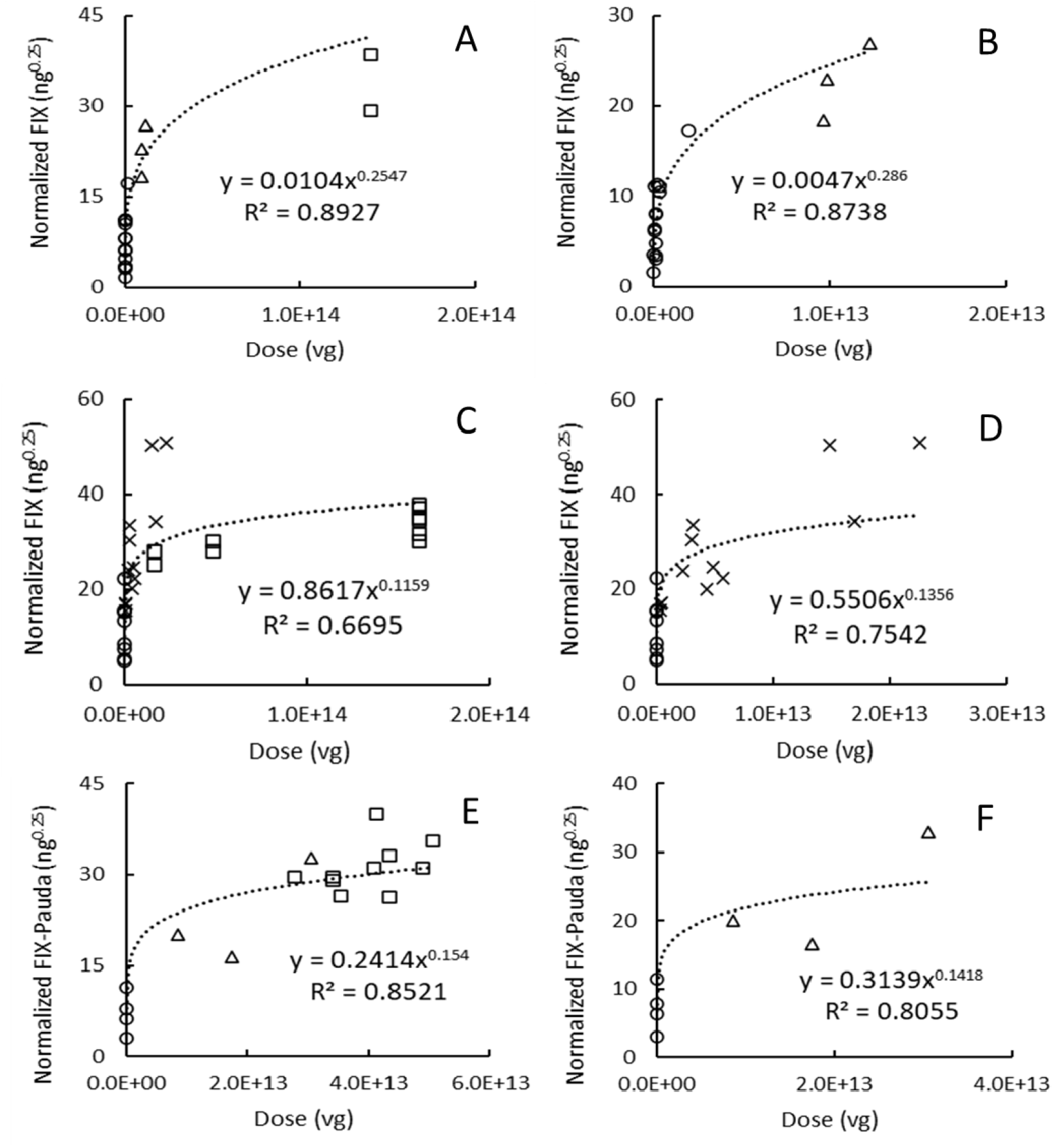
Normalized Factor IX-dose curves for (A and B) rAAV2-hAAT-FIX, (C and D) scAAV2/8-LP1-hFIXco, and (E and F) SPK-9001. The power regression was conducted using data from three species in A, C, and E and using data from two preclinical species in B, D, and F. Circle, triangle, crossing, and square symbols represent normalized FIX amounts in blood circulation of mouse, dog, macaque, and human, respectively.

**Figure 2.**
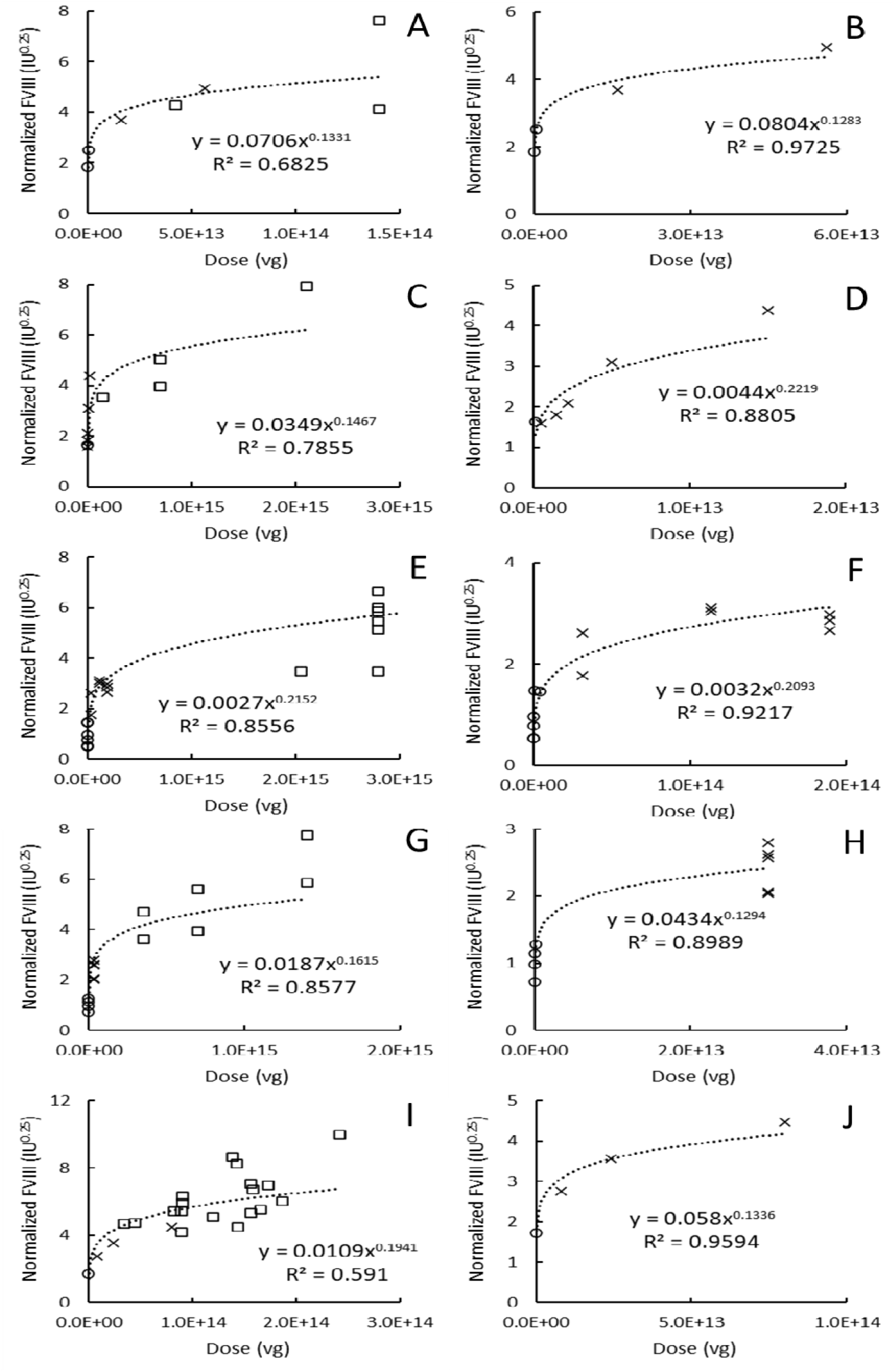
Normalized Factor VIII-dose curves for (A and B) GO-8, (C and D) SB-525, (E and F) BMN270, (G and H) DTX201, and (I and J) SPK-8011. The power regression was conducted using data from three species in A, C, E, G, and I and using data from two preclinical species in B, D, F, H, and J. Circle, triangle, crossing, and square symbols represent normalized FVIII amounts in blood circulation of mouse, dog, macaque, and human, respectively.

A power model was also used to generate regression equations based on the data of two preclinical species (mouse and dog or monkey) (Figures 1 and 2). The regression equations derived from preclinical data were then used to predict FIH doses of eight vectors which were anticipated to achieve targeted plasma levels of FIX (250 ng/mL), FIX-Pauda (27.5 ng/mL) or FVIII (12 IU/dL). As a comparison, FIH doses of the eight vectors were also predicted using direct vg/kg conversion and allometric scaling approaches.^5, 9^ The plots of allometric scaling analyses were shown in supplemental Figures S3 and S4. The FIH doses predicted by the three approaches were compared with clinical doses (Table 1). The FIH doses predicted by the three approaches were ranked as: allometric scaling > dose-response normalization > direct vg/kg conversion for most vectors except for SPK9001. For SPK9001, the predicted FIH doses were ranked as: allometric scaling > direct vg/kg conversion > dose-response normalization. Among the eight vectors, dose-response normalization generated more accurate FIH doses for three vectors (SPK-9001, GO-8, and BMN270). Allometric scaling provided more accurate predictions for three other vectors (rAAV2-hAAT-FIX, scAAV2/8-LP1-hFIXco and SB-525), and direct vg/kg conversion gave more accurate predictions for DTX201 and SPK-8011.

**Table 1.**
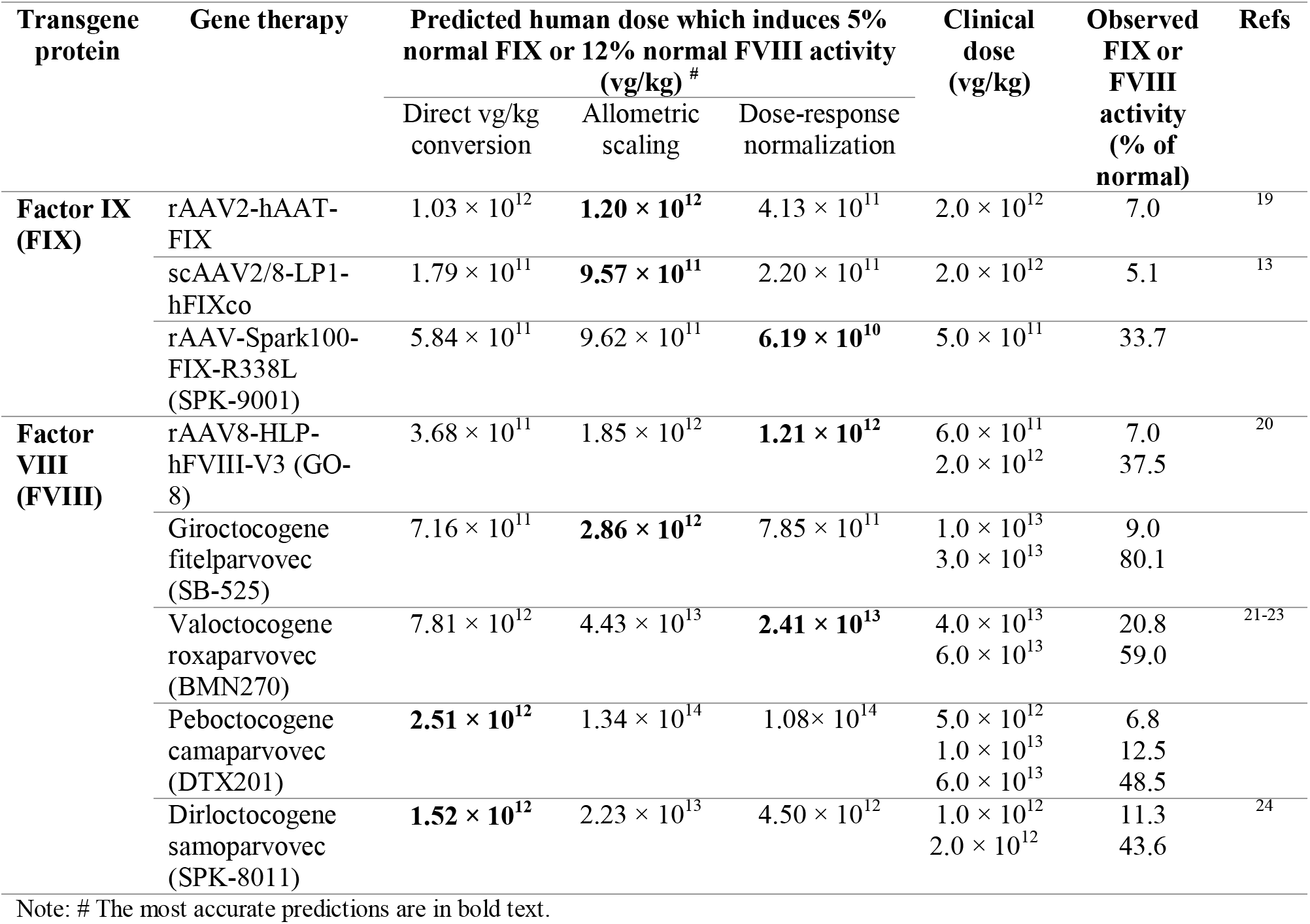
A comparison of interspecies normalization, allometric scaling, and direct vg/kg conversion approaches for human AAV vector dose predictions.

As shown in Figures 1 and 2, the total vector dose (vg) received by patients were usually much higher than that received by animals. To extrapolate dose-response from animals to patients, ideally, animal and human vector dose ranges should overlap. The animal and human dose ranges of scAAV2/8-LP1-hFIXco (Figure 1C), SPK-9001 (Figure 1E), GO-8 (Figure 2A), and SPK-8011 (Figure 2I) overlapped. Compared to direct vg/kg conversion and allometric scaling, dose-response normalization provided more accurate FIH dose predictions for SPK-9001 and GO-8 (Table 1). All the three approaches underestimated FIH dose of scAAV2/8-LP1-hFIXco and dose-response normalization underestimated the FIH dose by 9-fold, which was likely caused by intense T cell responses to the vector capsid in patients. Seven among 8 patients receiving medium-to-high dose of scAAV2/8-LP1-hFIXco reported T-cell responses against capsid, which could eliminate vector-transduced hepatocytes (Table S4).^13, 14^ All the three approaches overestimated FIH dose of SPK-8011. Dose-response normalization overestimated FIH dose of SPK-8011 by approximately 4-fold and direct vg/kg conversion provided a more accurate prediction. SPK-8011 contains bio-engineered AAV capsid which can transduce human hepatocytes more efficiently than animal hepatocytes. In a humanized mouse liver model, the bio-engineered SPK-8011 vector transduced primary human hepatocytes 100-fold higher than natural AAV8.^15^ The exceptionally high transduction efficiency of SPK8011 in human hepatocytes might lead to the overprediction of FIH dose.

For vectors rAAV2-hAAT-FIX, SB-525, BMN270, and DTX201, the dose ranges of animals and patients did not overlap. For example, the lowest patient dose and the highest monkey dose of SB-525 were 1.4 × 10^14^ vg and 1.5 × 10^13^ vg, respectively (Figure 2C and 2D). The non-overlapping pattern between animal and human doses might compromise animal-to-human extrapolation results. Among the four vectors, dose-response normalization provided an accurate prediction of FIH dose for BMN270 only. Similar to scAAV2/8-LP1-hFIXco, all the three approaches underestimated FIH dose of rAAV2-hAAT-FIX and dose-response normalization underestimated the FIH dose by 4 - 5 fold. Consistently, patients receiving a high dose of rAAV2-hAAT-FIX developed capsid-specific T cell responses (supplemental Table S4). In contrast, T-cell responses against capsid were either not observed or not investigated in the clinical studies of other vectors. The exact reasons for poor FIH dose predictions for SB-525 and DTX201 by dose-response normalization are not clear but may be associated with the non-overlapping pattern between human and animal doses.

## Discussion

Due to potential long-term safety risks, FIH study of most GT products is a dose-finding study conducted in a dozen or so patients with a rare disease of genetic disorders. Each patient usually receives a single dose of AAV vector only because the immune responses caused by repeated dosing may render subsequent administrations ineffective^9^. A patient receiving a low and ineffective starting dose may be ineligible to receive an escalated dose due to activated immunity against this AAV vector. Thus, an accurate prediction of FIH dose is critical for clinical development of GT products. In addition to the direct vg/kg conversion approach widely used in clinical development of AAV GT^16^ and recently reported allometric scaling approach,^5^ this study demonstrated that interspecies dose-response normalization could be used as an additional approach for FIH dose prediction for AAV-mediated GT. This new approach bypasses pharmacokinetic data and extrapolates dose-response relationship from preclinical species to hemophilia patients. This approach is especially useful at the early stage of clinical development when clinical data are insufficient to support the development and validation of complex quantitative systems pharmacology models. However, the performance of the interspecies dose-response normalization approach can be compromised by poor quality of preclinical data, uncertainties in human immune responses to vector/transgene product, and substantial interspecies differences in vector transduction efficiency and transgene expression efficiency.

The quality of preclinical data is critical for successfully extrapolating dose response from animals to humans. Preclinical dose-finding studies for AAV-mediated GT products are usually conducted in dogs or monkeys with a small sample size (N = 2 – 6 per dose level) and within a limited dose range. Due to the large inter-individual variability in transgene product levels, a preclinical dose-finding study with appropriate sample size is recommended. As shown in our regression analysis, the total vector dose range often did not overlap between dogs/monkeys and humans, which might compromise the quality of interspecies dose-response extrapolation. This issue can be addressed by (1) including large animal models with a body weight comparable to human (i.e., swine and cattle) into does-finding studies, and (2) testing GT products in the large animal models with a wide dose range. Since intracellular synthesis rate of transgene product is reversely correlated with W^0.25^, large animal models are anticipated to be more representative of humans in term of transgene product expression efficiency. The quality of both allometric scaling and interspecies dose-response normalization are expected to be improved by including dose response data of large animal models.^17^

Immune responses to vectors, transgene products, and AAV capsid antigens expressed on the surface of transduced cells in animals and humans are a big challenge in extrapolating dose-response from animals to humans.^8^ AAV capsid-specific T cell responses were detected in hemophilia B patients receiving moderate- or high-dose rAAV2-hAAT-FIX or scAAV2/8-LP1-hFIXco vectors. The presence of T-cell responses might eliminate vector-transduced human hepatocytes, decrease transgene product expression, and thus lead to substantial underprediction of FIH dose.^8^ Several steps may be taken to reduce the risk of AAV capsid-specific T cell responses: (1) develop appropriate in vitro tests (i.e., enzyme-linked immunospot assay and cytotoxic T lymphocyte assay) to identify and eliminate vector candidates with a high risk of T cell responses^16^; (2) exclude patients with preexisting immunity from FIH studies; and (3) maximize vector potency through capsid engineering and use high-activity transgene product variant and thus reduce FIH dose because AAV capsid-specific T cell responses are dose-dependent.^8, 18^

With the advance in capsid engineering and availability of new potent promoters/enhancers, some bioengineered AAV vectors showed a much higher transduction efficiency and a similar or even higher transgene expression efficiency in human hepatocytes than that in animal hepatocytes,^15^ which might cause an overprediction of FIH dose using allometric scaling or interspecies dose-response normalization approaches. The large interspecies differences in transduction efficiency and transgene expression efficiency can be detected using a humanized mouse liver model.^15^ These interspecies differences should be considered when dose response is extrapolated from animals to humans.

In this study, the proposed interspecies dose-response normalization was assessed with AAV vectors for hemophilia therapy only. Further work is needed to assess if this approach can be applied to (1) AAV-encoding transgene products that function in systemic circulation for other diseases; (2) other intravenously administered viral (e.g., adenoviral and lentiviral vectors) and non-viral GT modalities (e.g., lipid nanoparticles); and (3) locally administered GT products (e.g., intrathecal or intravitreal administration).

## Conclusions

In this study, the author first demonstrated that it was feasible to normalize dose response relationship of AAV-mediated hemophilia GT in multiple species to a species-invariant scale. The normalized dose response relationship from preclinical species could be extrapolated to hemophilia patients and used to predict FIH dose of AAV vectors for hemophilia therapy. In addition to hemophilia dogs and non-human primates, inclusion of larger animal models (e.g., swine and cattle) in preclinical dose-finding studies of AAV vectors is expected to improve the performance of interspecies dose-response normalization approach. Furthermore, AAV capsid-specific T cell responses in hemophilia patients may cause underprediction of FIH dose by the interspecies dose-response normalization approach. On the other hand, novel bioengineered capsids with a high transduction efficiency specifically in human hepatocytes may cause overprediction of FIH dose. Prior to a FIH study, in vitro tests should be used to detect AAV vector candidates with a high risk of AAV capsid-specific T cell responses and humanized animal liver models can be used to detect AAV vector candidates with a large interspecies difference in hepatocyte transduction. These interspecies differences in immune responses and vector transduction efficiency should be considered when dose response is extrapolated from preclinical species to patients.

## Data Availability

All data produced in the present work are contained in the manuscript.

## Conflicts of Interest

P.Z. is a current employee of Daiichi Sankyo Inc.

## Funding

This research received no specific grant from any funding agency in the public, commercial, or not-for-profit sectors.

## Data Sharing Statement

The data that support the findings of this study were collected from published literature and are summarized in supplementary material.

## Notes

### Competing Interest Statement

The authors have declared no competing interest.

### Funding Statement

This study did not receive any funding.

### Author Declarations

The study used ONLY openly available human data collected from literature.

