## Supplementary materials for "Interspecies normalization of dose response relationship for adeno-associated virus-mediated hemophilia gene therapy – application to first-in-human dose prediction"

Peng Zou, Ph.D.*

Quantitative Clinical Pharmacology, Daiichi Sankyo, Inc., 211 Mt. Airy Road, Basking Ridge, NJ 07920

*Corresponding author

Peng Zou, Ph.D.

Quantitative Clinical Pharmacology

Daiichi Sankyo, Inc.

211 Mount Airy Road

Basking Ridge, NJ 07920

**Supplemental Figure S1. Normalized mean Factor IX-dose curves for (A and B) rAAV2-hAAT-FIX, (C and D) scAAV2/8-LP1-hFIXco, and (E and F) rAAV-Spark100-FIX-R338L.** The power regression was conducted using data from three species in A, C, and E and using data from two preclinical species in B, D, and F. Circle, triangle, crossing, and square symbols represent normalized mean FIX amounts in blood circulation of mouse, dog, macaque, and human, respectively.

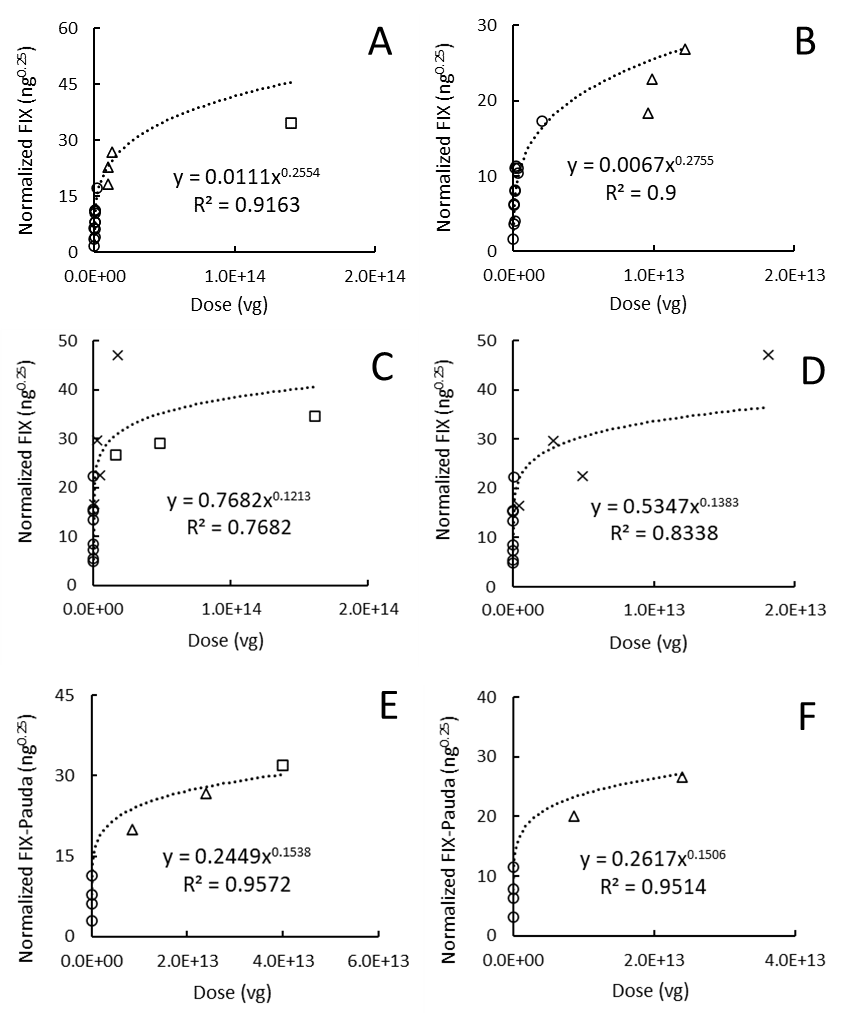

**Supplemental Figure S2. Normalized mean Factor VIII-dose curves for (A and B) GO-8, (C and D) SB-525, (E and F) BMN270, (G and H) DTX201, and (I and J) SPK-8011.** The power regression was conducted using data from three species in A, C, E, G, and I and using data from two preclinical species in B, D, F, H, and J. Circle, triangle, crossing, and square symbols represent normalized mean FVIII amounts in blood circulation of mouse, dog, macaque, and human, respectively.

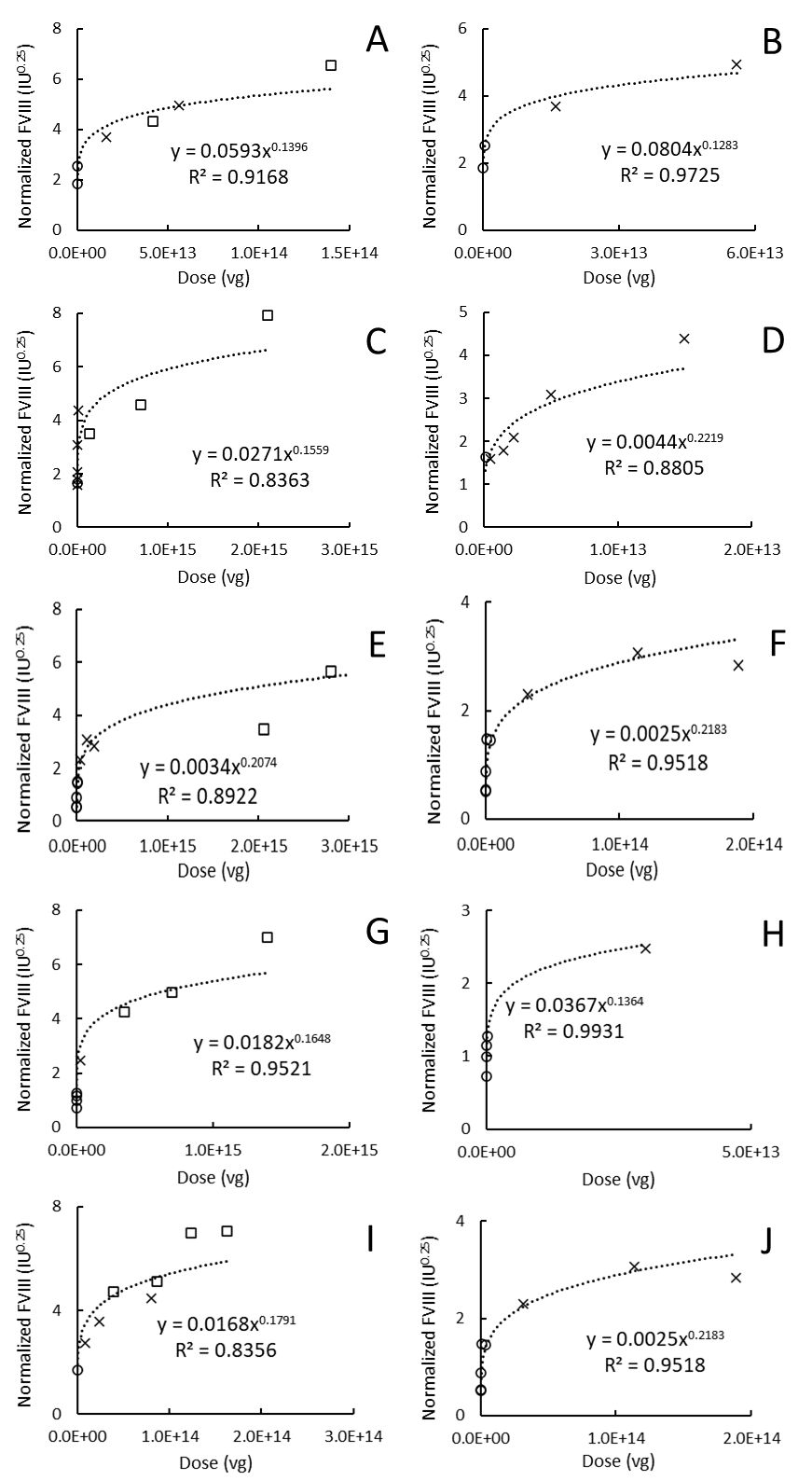

**Supplemental Figure S3. Allometric scaling of gene efficiency factor (GEF) for (A and B) rAAV2-hAAT-FIX, (C and D) scAAV2/8-LP1-hFIXco, and (E and F) SPK-9001.** Allometric scaling was conducted using data from three species in A, C, and E and using data from two preclinical species in B, D, and F. LogW was used as the scaling variable. W is body weight in kg and the unit of GEF is molecules/day/viral genome. Circle, triangle, square, and diamond symbols represent mouse, dog, macaque, and human respectively.

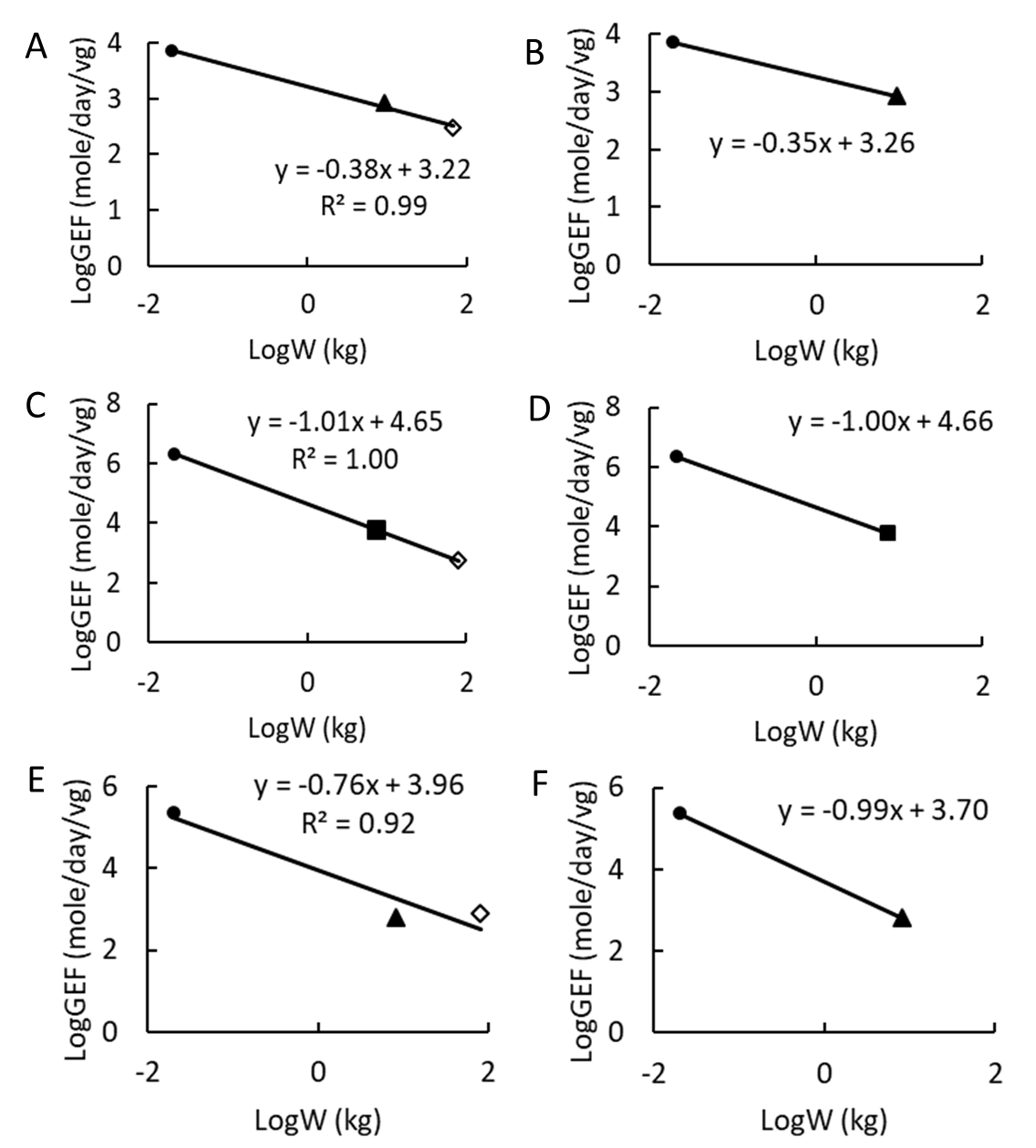

**Supplemental Figure S4. Allometric scaling of gene efficiency factor (GEF) for (A and B) GO-8, (C and D) SB-525, (E and F) BMN270, (G and H) DTX201, and (I and J) SPK-8011**. Allometric scaling was conducted using data from three species in A, C, E, G, and I and using data from two preclinical species in B, D, F, H, and J. LogW was used as the scaling variable. W is body weight in kg and the unit of GEF is IU/day/viral genome. Circle, triangle, square, and diamond symbols represent mouse, dog, macaque, and human respectively.

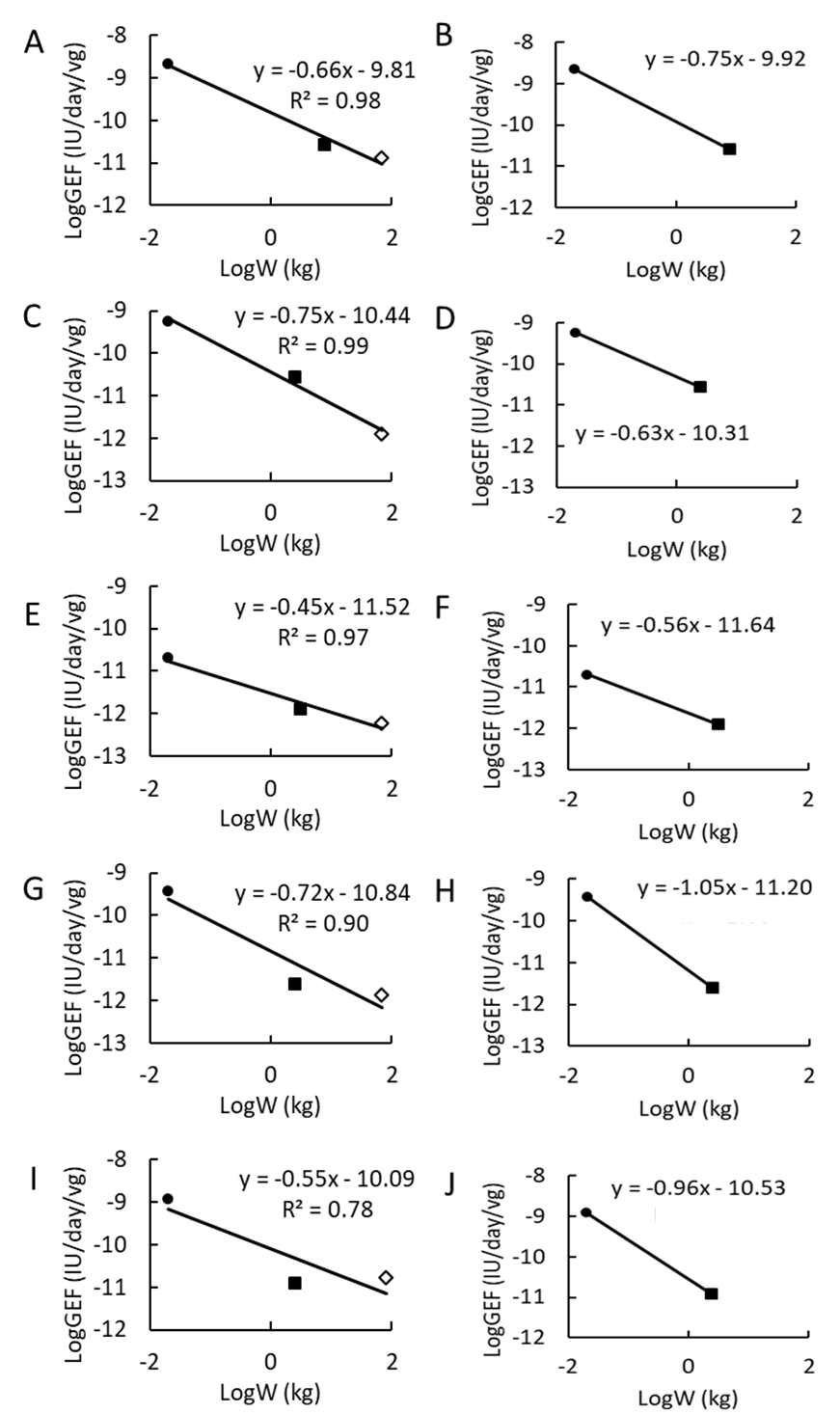

**Supplemental Table S1. Clearance values of recombinant factor IX and factor VIII used for gene efficiency factor calculations.**

| Species | CL_FIX_ (mL/h/kg) | CL_FVIII_ (mL/h/kg) |
| --- | --- | --- |
| Mice | 34.8 ± 3.1 ^1^ | 37.83 ± 36.1 ^2-4^ |
| Dogs | 9.96 ± 0.55 ^1^ | N.A. |
| Monkeys | 8.42 ± 2.00 ^1^ | 5.21 ^4^ |
| Humans | 7.58 ± 1.84 ^1^ | 3.30 ± 0.28 ^2, 5^ |

**Supplemental Table S2. Plasma FVIII activity, gene efficiency factor, and normalized FVIII values used for regression analysis.**

| Vector | Species | Dose (vg/kg) | Body weight (kg) | Blood volume (mL/kg) | Plasma FVIII activity (IU/dL or % of normal) | Normalized FVIII in blood circulation (IU^0.25^) | GEF (IU/day/vg) | Refs |
| --- | --- | --- | --- | --- | --- | --- | --- | --- |
| rAAV8-HLP-hFVIII-V3 (GO-8) | Mice | 2.00E+12 | 0.02 | 79 | 732 | 1.84 | 3.32E-09 | ^6^ |
|  |  | 2.00E+13 | 0.02 |  | 2578 | 2.53 | 1.17E-09 |  |
|  | Monkeys (Rhesus) | 2.00E+12 | 8 | 54 | 43 | 3.69 | 2.69E-11 | ^6^ |
|  |  | 7.00E+12 | 8 |  | 138 | 4.94 | 2.47E-11 |  |
|  | Humans | 6.00E+11 | 70 | 70 | 7 | 4.30 | 9.24E-12 | ^7^ |
|  |  | 2.00E+12 | 70 |  | 6 | 4.14 | 2.38E-12 |  |
|  |  | 2.00E+12 | 70 |  | 69 | 7.63 | 2.73E-11 |  |
| SB-525 | Mice | 7.20E+12 | 0.02 | 79 | 458.1 | 1.64 | 5.78E-10 | ^8^ |
|  | Monkeys (Cynomolgus) | 2.00E+11 | 2.5 | 65 | 3.9 | 1.59 | 2.44E-11 | ^8^ |
|  |  | 6.00E+11 | 2.5 |  | 6.4 | 1.80 | 1.33E-11 |  |
|  |  | 9.00E+11 | 2.5 |  | 11.7 | 2.09 | 1.63E-11 |  |
|  |  | 2.00E+12 | 2.5 |  | 56.4 | 3.09 | 3.53E-11 |  |
|  |  | 6.00E+12 | 2.5 |  | 227.9 | 4.39 | 4.75E-11 |  |
|  | Humans | 2.00E+12 | 70 | Human | 3.1 | 3.51 | 1.23E-12 | ^9^ |
|  |  | 1.00E+13 | 70 |  | 5 | 3.96 | 3.96E-13 |  |
|  |  | 1.00E+13 | 70 |  | 13 | 5.02 | 1.03E-12 |  |
|  |  | 3.00E+13 | 70 |  | 80.1 | 7.92 | 2.11E-12 |  |
| BMN270 | Mice | 2.00E+12 | 0.02 | 79 | 4.69 | 0.52 | 2.13E-11 | ^10^ |
|  |  | 2.00E+13 | 0.02 |  | 23.5 | 0.78 | 1.07E-11 |  |
|  |  | 2.00E+14 | 0.02 |  | 287 | 1.46 | 1.30E-11 |  |
|  |  | 6.00E+12 | 0.02 |  | 4.9 | 0.53 | 7.41E-12 |  |
|  |  | 2.00E+13 | 0.02 |  | 53 | 0.96 | 2.41E-11 |  |
|  |  | 6.00E+13 | 0.02 |  | 299 | 1.47 | 4.52E-11 |  |
|  | Monkeys (Cynomolgus) | 1.00E+13 | 3.15 | 65 | 22.8 | 2.61 | 2.85E-12 | ^10, 11^ |
|  |  | 1.00E+13 | 3.15 |  | 4.8 | 1.77 | 6.00E-13 |  |
|  |  | 3.60E+13 | 3.15 |  | 41.3 | 3.03 | 1.43E-12 |  |
|  |  | 3.6E+13 | 3.15 |  | 45.6 | 3.11 | 1.58E-12 |  |
|  |  | 6.00E+13 | 3.15 |  | 32.9 | 2.86 | 6.86E-13 |  |
|  |  | 6.00E+13 | 3.15 |  | 38.2 | 2.97 | 7.96E-13 |  |
|  |  | 6.00E+13 | 3.15 |  | 24.3 | 2.66 | 5.06E-13 |  |
|  | Humans | 2.00E+13 | 103 | 70 | 2 | 3.47 | 7.92E-14 | ^12-14^ |
|  |  | 4.00E+13 | 70 |  | 18 | 5.45 | 3.56E-13 |  |
|  |  | 4.00E+13 | 70 |  | 40 | 6.65 | 7.92E-13 |  |
|  |  | 4.00E+13 | 70 |  | 24 | 5.86 | 4.75E-13 |  |
|  |  | 4.00E+13 | 70 |  | 14 | 5.12 | 2.77E-13 |  |
|  |  | 4.00E+13 | 70 |  | 26 | 5.97 | 5.15E-13 |  |
|  |  | 4.00E+13 | 70 |  | 3 | 3.48 | 5.94E-14 |  |
|  |  | 6.00E+13 | 70 |  | 62 | 7.42 | 8.18E-13 |  |
|  |  | 6.00E+13 | 70 |  | 50 | 7.04 | 6.60E-13 |  |
|  |  | 6.00E+13 | 70 |  | 95 | 8.26 | 1.25E-12 |  |
|  |  | 6.00E+13 | 70 |  | 11 | 4.82 | 1.45E-13 |  |
|  |  | 6.00E+13 | 70 |  | 88 | 8.10 | 1.16E-12 |  |
|  |  | 6.00E+13 | 70 |  | 52 | 7.10 | 6.86E-13 |  |
|  |  | 6.00E+13 | 70 |  | 55 | 7.21 | 7.26E-13 |  |
| DTX201 | Mice | 3.00E+11 | 0.02 | 79 | 17.3 | 0.72 | 5.24E-10 | ^15^ |
|  |  | 1.00E+12 | 0.02 |  | 60 | 0.99 | 5.45E-10 |  |
|  |  | 3.00E+12 | 0.02 |  | 110 | 1.15 | 3.33E-10 |  |
|  |  | 1.00E+13 | 0.02 |  | 168.4 | 1.28 | 1.53E-10 |  |
|  | Monkeys (Cynomolgus) | 1.20E+13 | 2.5 | 65 | 37.1 | 2.79 | 3.87E-12 | ^16^ |
|  |  | 1.20E+13 | 2.5 |  | 10.5 | 2.03 | 1.09E-12 |  |
|  |  | 1.20E+13 | 2.5 |  | 11 | 2.06 | 1.15E-12 |  |
|  |  | 1.20E+13 | 2.5 |  | 27 | 2.57 | 2.81E-12 |  |
|  |  | 1.20E+13 | 2.5 |  | 29 | 2.62 | 3.02E-12 |  |
|  | Humans | 5.00E+12 | 70 | 70 | 10 | 4.70 | 1.58E-12 | ^17^ |
|  |  | 5.00E+12 | 70 |  | 3.5 | 3.62 | 5.54E-13 |  |
|  |  | 1.00E+13 | 70 |  | 20 | 5.60 | 1.58E-12 |  |
|  |  | 1.00E+13 | 70 |  | 5 | 3.96 | 3.96E-13 |  |
|  |  | 2.00E+13 | 70 |  | 73 | 7.73 | 2.89E-12 |  |
|  |  | 2.00E+13 | 70 |  | 24 | 5.86 | 9.50E-13 |  |
| SPK-8011 | Mice | 4.00E+12 | 0.02 | 79 | 550 | 1.72 | 1.25E-09 | ^18^ |
|  | Monkeys (Cynomolgus) | 2.00E+12 | 4 | 65 | 22.3 | 2.76 | 1.39E-11 | ^18^ |
|  |  | 6.00E+12 | 4 |  | 62 | 3.56 | 1.29E-11 |  |
|  |  | 2.00E+13 | 4 |  | 153 | 4.47 | 9.57E-12 |  |
|  | Humans | 5.00E+11 | 68 | 70 | 10 | 4.67 | 1.58E-11 | ^19^ |
|  |  | 5.00E+11 | 89 |  | 8 | 4.72 | 1.27E-11 |  |
|  |  | 1.00E+12 | 89 |  | 5 | 4.20 | 3.96E-12 |  |
|  |  | 1.00E+12 | 82 |  | 15 | 5.42 | 1.19E-11 |  |
|  |  | 1.00E+12 | 89 |  | 14 | 5.43 | 1.11E-11 |  |
|  |  | 2.00E+12 | 60 |  | 16 | 5.09 | 6.34E-12 |  |
|  |  | 2.00E+12 | 79 |  | 37 | 6.73 | 1.47E-11 |  |
|  |  | 2.00E+12 | 93 |  | 20 | 6.01 | 7.92E-12 |  |
|  |  | 2.00E+12 | 72 |  | 8 | 4.48 | 3.17E-12 |  |
|  |  | 2.00E+12 | 78 |  | 15 | 5.35 | 5.94E-12 |  |
|  |  | 2.00E+12 | 78 |  | 46 | 7.08 | 1.82E-11 |  |
|  |  | 2.00E+12 | 83 |  | 16 | 5.52 | 6.34E-12 |  |
|  |  | 2.00E+12 | 69 |  | 117 | 8.67 | 4.63E-11 |  |
|  |  | 2.00E+12 | 121 |  | 117 | 9.98 | 4.63E-11 |  |
|  |  | 1.50E+12 | 60 |  | 29 | 5.91 | 1.53E-11 |  |
|  |  | 1.50E+12 | 115 |  | 29 | 6.95 | 1.53E-11 |  |
|  |  | 1.50E+12 | 60 |  | 38 | 6.32 | 2.01E-11 |  |
|  |  | 1.50E+12 | 95 |  | 70 | 8.26 | 3.70E-11 |  |

**Supplemental Table S3. Plasma Factor IX levels, gene efficiency factor, and normalized Factor IX values used for regression analysis.**

| Vector | Species | Dose (vg/kg) | Body weight (kg) | Blood volume  (mL/kg) | Plasma FIX (ng/mL) | Normalized FIX in blood circulation (ng^0.25^) | GEF (molecules/day/vg) | Refs |
| --- | --- | --- | --- | --- | --- | --- | --- | --- |
| rAAV2-hAAT-FIX | Mice | 1.67E+13 | 0.018 | 79 | 8410 | 10.46 | 4610 | ^1^ |
|  |  | 4.00E+12 | 0.02 |  | 9630 | 11.11 | 22000 |  |
|  |  | 1.04E+12 | 0.0193 |  | 1060 | 6.34 | 9360 |  |
|  |  | 1.04E+13 | 0.0193 |  | 10900 | 11.35 | 9630 |  |
|  |  | 1.04E+14 | 0.0193 |  | 58500 | 17.28 | 5160 |  |
|  |  | 5.18E+12 | 0.0193 |  | 2750 | 8.05 | 4850 |  |
|  |  | 1.85E+11 | 0.02 |  | 4.07 | 1.59 | 201 |  |
|  |  | 5.50E+11 | 0.02 |  | 103 | 3.57 | 1710 |  |
|  |  | 1.65E+12 | 0.02 |  | 897 | 6.14 | 4970 |  |
|  |  | 5.50E+12 | 0.02 |  | 2820 | 8.17 | 5160 |  |
|  |  | 1.50E+13 | 0.02 |  | 9550 | 11.08 | 5820 |  |
|  |  | 5.00E+12 | 0.02 |  | 350 | 4.85 | 639 |  |
|  |  | 5.00E+12 | 0.02 |  | 87.1 | 3.43 | 159 |  |
|  |  | 5.00E+12 | 0.02 |  | 60.9 | 3.13 | 111 |  |
|  | Dogs | 1.20E+12 | 10.2 | 86 | 590 | 26.82 | 1260 | ^1^ |
|  |  | 1.60E+12 | 6 |  | 220 | 18.36 | 356 |  |
|  |  | 8.00E+11 | 12.3 |  | 262 | 22.94 | 878 |  |
|  | Humans | 2.00E+12 | 70 | 70 | 444 | 38.41 | 442 | ^20^ |
|  |  | 2.00E+12 | 70 |  | 150 | 29.28 | 149* |  |
| scAAV2/8-LP1-hFIXco | Mice | 5.00E+10 | 0.02 | 79 | 563 | 5.46 | 103000 | ^1^ |
|  |  | 1E+11 | 0.02 |  | 3250 | 8.47 | 297000 |  |
|  |  | 1.25E+12 | 0.02 |  | 36600 | 15.51 | 268000 |  |
|  |  | 5E+12 | 0.02 |  | 155000 | 22.25 | 284000 |  |
|  |  | 2.00E+11 | 0.0227 |  | 30400 | 15.28 | 1390000 |  |
|  |  | 4.00E+10 | 0.0227 |  | 17700 | 13.35 | 4040000 |  |
|  |  | 4.00E+09 | 0.0227 |  | 1610 | 7.33 | 3680000 |  |
|  |  | 4.00E+08 | 0.0227 |  | 322 | 4.90 | 7350000 |  |
|  | Monkeys (Cynomolgus) | 1.00E+12 | 4.9 | 65 | 1400 | 24.67 | 3100 | ^1^ |
|  |  | 1.00E+12 | 5.7 |  | 800 | 22.28 | 1770 |  |
|  |  | 1.00E+12 | 4.3 |  | 700 | 20.08 | 1550 |  |
|  |  | 2.00E+12 | 8.5 |  | 3000 | 34.26 | 3320 |  |
|  |  | 2.00E+12 | 7.4 |  | 16100 | 50.36 | 17900 |  |
|  |  | 2.00E+12 | 11.3 |  | 11000 | 50.89 | 12200 |  |
|  |  | 2.00E+11 | 11.7 |  | 523 | 23.98 | 5780 |  |
|  |  | 2.00E+11 | 15.1 |  | 1030 | 30.27 | 14400 |  |
|  |  | 2.00E+11 | 15.7 |  | 1490 | 33.53 | 16400 |  |
|  |  | 6.00E+10 | 6.7 |  | 161 | 15.54 | 5940 |  |
|  |  | 6.00E+10 | 7.3 |  | 213 | 17.02 | 7850 |  |
|  |  | 6.00E+10 | 6.3 |  | 235 | 16.82 | 8660 |  |
|  | Humans | 2.00E+11 | 80.7 | 70 | 109 | 28.01 | 889 | ^1^ |
|  |  | 2.00E+11 | 80.7 |  | 70 | 25.08 |  |  |
|  |  | 6.00E+11 | 80.7 |  | 143 | 29.98 | 417 |  |
|  |  | 6.00E+11 | 80.7 |  | 109 | 28.01 |  |  |
|  |  | 2.00E+12 | 80.7 |  | 178 | 31.67 | 254 |  |
|  |  | 2.00E+12 | 80.7 |  | 361 | 37.79 |  |  |
|  |  | 2.00E+12 | 80.7 |  | 250 | 34.47 |  |  |
|  |  | 2.00E+12 | 80.7 |  | 334 | 37.06 |  |  |
|  |  | 2.00E+12 | 80.7 |  | 262 | 34.88 |  |  |
|  |  | 2.00E+12 | 80.7 |  | 145 | 30.08 |  |  |
| rAAV-Spark100-FIX-R338L (SPK-9001) | Mice | 1.00E+10 | 0.02 | 79 | 53.8 | 3.04 | 270180 | ^21^ |
|  |  | 4.00E+10 | 0.02 |  | 173.5 | 6.23 | 217826 |  |
|  |  | 1.00E+11 | 0.02 |  | 424.1 | 7.79 | 212980 |  |
|  |  | 4.00E+11 | 0.02 |  | 1978.5 | 11.45 | 248398 |  |
|  | Dogs | 1.00E+12 | 8.6 | 86 | 39.9 | 20.07 | 573 | ^21^ |
|  |  | 3.00E+12 | 10.2 |  | 241.2 | 32.84 | 1156 |  |
|  |  | 3.00E+12 | 5.8 |  | 26.8 | 16.46 | 128 |  |
|  | Humans | 5.00E+11 | 81.8 | 70 | 295.6 | 31.17 | 656 | ^22^ |
|  |  | 5.00E+11 | 55.6 |  | 953.3 | 29.62 | 788 |  |
|  |  | 5.00E+11 | 97.9 |  | 2330.2 | 31.15 | 547 |  |
|  |  | 5.00E+11 | 101.3 |  | 10870.9 | 35.55 | 897 |  |
|  |  | 5.00E+11 | 87 |  | 219.2 | 33.13 | 788 |  |
|  |  | 5.00E+11 | 70.9 |  | 1325.3 | 26.47 | 394 |  |
|  |  | 5.00E+11 | 87 |  | 147.3 | 26.16 | 306 |  |
|  |  | 5.00E+11 | 68.2 |  | 164.8 | 29.01 | 591 |  |
|  |  | 5.00E+11 | 82.8 |  | 197.8 | 40.08 | 1772 |  |
|  |  | 5.00E+11 | 68.1 |  | 137.4 | 29.52 | 634 |  |

*Subject F with plasma FIX level of 150 ng/mL (3% of normal level) was included in the calculation ^20^.

**Supplemental Table S4. Clinical immunogenicity data of eight vectors by dose cohort**

| Gene therapy | Route of administration | Dose (vg/kg) | Number of patients | Safety data | Refs |
| --- | --- | --- | --- | --- | --- |
| rAAV2-hAAT-FIX | IV infusion | 8x10^10^ | 2 | Increase in ALT and AST (n=2)  **Two patients in the high dose cohort developed T-cell response against capsid**. | ^20, 23^ |
|  |  | 4x10^11^ | 3 |  |  |
|  |  | 2x10^12^ | 2 |  |  |
| scAAV2/8-LP1-hFIXco | IV infusion | 2x10^11^ | 2 | Increased AAV8-IgG antibody titers (n=2) | ^24, 25^ |
|  |  | 6x10^11^ | 2 | **T-cell response against capsid (n=2),** Increased AAV8-IgG antibody titers (n=2) |  |
|  |  | 2x10^12^ | 6 | **T-cell response against capsid (n=5),** elevation in ALT (n=4), Increased AAV8-IgG antibody titers (n=6) |  |
| SPK-9001 | IV infusion | 6x10^11^ | 10 | Increase in ALT (n=2) | ^22^ |
| GO-8 | IV infusion | 6x10^11^ | 1 | Increase in ALT (n=2) | ^26^ |
|  |  | 2x10^12^ | 2 |  |  |
| SB-525 |  | 9x10^11^ | 2 | Increase in ALT (n=4) | ^9^ |
|  |  | 2x10^12^ | 2 |  |  |
|  |  | 1x10^13^ | 2 |  |  |
|  |  | 3x10^13^ | 5 | Increase in ALT (n=4) |  |
| BMN270 | IV infusion | 6x10^12^ | 1 | Increase in ALT and anti-AAV5 antibodies (n=1) | ^14^ |
|  |  | 2x10^13^ | 1 | Anti-AAV5 antibodies(n=1) |  |
|  |  | 6x10^13^ | 7 | Increase in ALT and anti-AAV5 antibodies (n=7) |  |
| DTX201 | IV infusion | 5x10^12^ | 2 | N.A. | ^17^ |
|  |  | 1x10^13^ | 2 | Increase in ALT and AST (n=1) |  |
|  |  | 2x10^13^ | 4 | Increase in ALT and AST (n=2) |  |
| SPK-8011 | IV infusion | 5x10^11^ | 2 | None | ^19^ |
|  |  | 1x10^12^ | 3 | None |  |
|  |  | 1.5x10^12^ | 4 | Increase in ALT (n=4) |  |
|  |  | 2x10^12^ | 9 | Increase in ALT (n=3) |  |
